# *CFAP47* is a novel causative gene implicated in X-linked polycystic kidney disease

**DOI:** 10.1101/2024.04.05.24304760

**Authors:** Takayasu Mori, Takuya Fujimaru, Chunyu Liu, Karynne Patterson, Kohei Yamamoto, Takefumi Suzuki, Motoko Chiga, Akinari Sekine, Yoshifumi Ubara, Danny E Miller, Miranda PG Zalusky, Shintaro Mandai, Fumiaki Ando, Yutaro Mori, Hiroaki Kikuchi, Koichiro Susa, University of Washington Center for Rare Disease Research, Jessica X. Chong, Michael J. Bamshad, Yue-Qiu Tan, Feng Zhang, Shinichi Uchida, Eisei Sohara

## Abstract

Autosomal dominant polycystic kidney disease (ADPKD) is a well-described condition in which ∼80% of cases have a genetic explanation, while the genetic basis of sporadic cystic kidney disease in adults remains unclear in ∼30% of cases. This study aimed to identify novel genes associated with polycystic kidney disease (PKD) in patients with sporadic cystic kidney disease in which a clear genetic change was not identified in established genes. A next-generation sequencing panel analyzed known genes related to renal cysts in 118 sporadic cases, followed by whole-genome sequencing on 47 unrelated individuals without identified candidate variants. Three male patients were found to have rare missense variants in the X-linked gene Cilia And Flagella Associated Protein 47 (*CFAP47*). CFAP47 was expressed in primary cilia of human renal tubules, and knockout mice exhibited vacuolation of tubular cells and tubular dilation, providing evidence that *CFAP47* is a causative gene involved in cyst formation. This discovery of *CFAP47* as a newly identified gene associated with PKD, displaying X-linked inheritance, emphasizes the need for further cases to understand the role of *CFAP47* in PKD.

## Introduction

Autosomal dominant polycystic kidney disease (ADPKD) is an autosomal dominant kidney disorder is marked by distinctive imaging features and is associated with mutations primarily in two genes, *PKD1* and *PKD2*. Typically manifesting in adulthood, the disease is characterized by advancing cyst formation and declining renal function. While genetic diagnostic rates vary among studies and centers, *PKD1* mutations are found in about 85-90% of ADPKD patients and *PKD2* mutations in about 10-15%^1^. One study identified the responsible mutation in 81% of patients diagnosed with clinically typical ADPKD^2^ and when ADPKD was studied in a large unselected cohort, only 77% of patients diagnosed with ADPKD in their medical records were identified with variants associated with the disease^3^. In a large ADPKD cohort studied in Taiwan, pathogenic variants in *PKD1* or *PKD2* were detected in only 69% of families, 7% of which were due to variants in genes other than *PKD1* and *PKD2* ^4^. Collectively, these observations suggest that a subset of cases are not explained by mutations in *PKD1* and *PKD2*.

Recent advances in genetic analysis technology have led to the identification of several new genes associated with ADPKD, including *GANAB* ^5^, *IFT140*^6^, *ALG5*^7^, *ALG9*, *DNAJB11*, and *HNF1B*, collectively explaining approximately 7% cases of ADPKD^8, 9^. In order to identify additional genes associated with this condition we conducted whole-genome sequencing (WGS) on 47 unrelated patients exhibiting solitary multiple renal cysts, where no responsible mutation was identified through a panel-based genetic screen for cystic kidney disease. We identified putatively pathogenic variants in the gene, *CFAP47*, encoding cilia and flagella-associated protein 47 (MIM: 301057), suggesting that it is a novel gene underlying cystic kidney disease.

## Materials and Methods

### Patients and ethics statement

This research received approval from the Institutional Review Board of Tokyo Medical and Dental University (approval number #G2000-081 and #M2019-324) and was conducted in accordance with the Declaration of Helsinki. All participants in the study provided written informed consent and agreed to the utilization of their DNA and kidney tissues in research aimed at identifying genetic risk variants for kidney function. Additionally, all participants consented to the publication of their genetic and medical data in academic journals, provided that the data were anonymized. The patient IDs (PT or K numbers) mentioned within the text are shared exclusively within the research group and are unknown to anyone else.

This is a multicenter cohort study. From 2014 to 2020, patients with sporadic polycystic kidney disease were recruited from 27 Japanese institutions, including Tokyo Medical and Dental University. Polycystic kidney disease was defined as having more than five cysts in each kidney on both sides, as detected by CT or MRI. In assessing family history, the presence or absence of cystic kidney disease in the parents and siblings was carefully determined based on their medical history, including previous illnesses and hospital visits. Adults aged 20 years and older were included in the study. Clinical data was gathered from medical records. The estimated glomerular filtration rate (eGFR) was calculated using the Japanese glomerular filtration rate equation^10^. Total kidney volume (TKV) was calculated from CT or MRI based on the volume of a modified ellipse for each kidney using the formula: volume = π / 6 × length × width × depth^11^.

### Genetic analysis

#### Targeted Next-Generation Sequencing (NGS) Panel Screening

Comprehensive genetic testing was performed using a capture-based targeted NGS method. We examined 69 genes (panel version 1) or 92 genes (panel version 2) associated with various inherited renal cystic diseases (refer to Supplementary table S1). The detailed methods are outlined in the Supplementary Methods and our prior publications^12, 13^. Following the initial filtering of variants based on minor allele frequencies (MAF), all variants underwent evaluation according to the American College of Medical Genetics and Genomics/Association for Molecular Pathology (ACMG/AMP) guidelines^14^. Variants categorized as “pathogenic” or “likely-pathogenic” were treated as pathogenic variants. To identify large genomic rearrangements, such as gross deletions or duplications, copy number variation analysis was conducted using Copy Number Analysis for Targeted Resequencing (CONTRA) ^15^.

#### Targeted long-read sequencing (T-LRS)

Prior to sequencing DNA was sheared with a g-TUBE (Covaris, Woburn, Massachusetts) at 6,000 rpm 2x. Approximately 1.5 ug of DNA was used for each library. Libraries were made using the Nanopore ligation kit (LSK-109) following their standard protocol. A single library was loaded onto a R9.4.1 flow cell on a GridION (Oxford Nanopore Technologies, Oxford, United Kingdom) and run for 72 hours. Targeted sequencing was done using Read-Until (Oxford Nanopore Technologies). Target regions are provided in Supplemental table S2. Reads were base called with Guppy 5.0.7 (Oxford Nanopore Technologies) using the super accurate model and aligned to GRCh38 with minimap2^16^. Variants were called and phased using Medaka (Oxford Nanopore Technologies), then annotated with VEP^17^ to include CADD^18^, SpliceAI^19^ scores, and gnomAD v3 allele frequency. SVs were called using Sniffles^20^ and SVIM^21^.

#### Whole genome sequencing analysis

Sequencing was performed at the University of Washington Center for Rare Disease Research (UW-CRDR). The detailed methods are outlined in the Supplementary Methods. Variant calling was performed using the HaplotypeCaller (HC) tool from GATK (3.7) ^22^. Variant filtering was conducted using seqr, a web-based Whole Genome Sequencing (WGS) analysis tool developed by the Broad Institute^23^. The analysis was generally limited to exonic and splice regions. Variants with a MAF in gnomAD ^24^ greater than 0.005 were excluded, and variants with a CADD score^18^ exceeding 18 or null (e.g. indels) were selected. As this study involved a cohort analysis of isolated cases, candidate variants were further refined based on autosomal recessive inheritance patterns (homozygous or presumed compound heterozygous variants) and X-chromosomal recessive inheritance mode.

### Animal experiments

All animal studies were carried out in accordance with the recommendations of the US National Institutes of Health’s Guide for the Care and Use of Laboratory Animals and the guidelines for animal research of Tokyo Medical and Dental University. The study was approved by the animal ethics committee at the School of Life Sciences of Fudan University and the Animal Care and Use Committee of Tokyo Medical and Dental University (approval number: A2023-109A). The investigators took all necessary measures to minimize the pain and suffering of the animals throughout the study. All mice were maintained under standard lightning conditions (12/12 h light/dark cycle). *Cfap47*-KO mice (*Cfap47^-/Y^*) carrying frameshift mutation^25^ generated by CRISPR-Cas9 technology were kindly provided by Feng Zhang laboratory in Fudan University, Shanghai, China. The strain background is C57BL/6. Adult mice (aged 40 weeks or older) were used in this study.

### Immunofluorescence studies

Formalin-fixed, paraffin-embedded specimens of normal human renal tissue are prepared. Heat-induced antigen retrieval was performed in a steamer using citrate buffer (H-3300; Vector Laboratories, Burlingame, CA) for 30 minutes. After being blocked with 1% bovine serum albumin, serial sections were then incubated with the primary antibody, mouse monoclonal anti-acetylated tubulin (1:5000; # T7451, MilliporeSigma) and rabbit anti-CXorf22 (Cfap47) polyclonal antibody (1:100, # 20837-1-AP, Proteintech Group, Inc.), for 2 hours at room temperature. The secondary antibodies utilized were Goat anti-Rabbit IgG (H+L) Cross-Adsorbed Secondary Antibody, Alexa Fluor™ 488 (#AB_143165), and 546 (#AB_2534071) (Invitrogen, Waltham, Massachusetts).

### Histological examination of mouse kidney

Kidney tissues were fixed with 4% paraformaldehyde, embedded in paraffin, and sectioned at 4 μm. Hematoxylin-eosin staining was conducted using standard procedures.

### Statistical analysis

Statistical significance was evaluated using unpaired t-tests. All data are presented as the mean ± standard deviation, and a P-value of < 0.05 was considered to be statistically significant.

## Results

### Patient Enrollment and Comprehensive Genetic Analysis

Between 2014 and 2020, we enrolled patients with sporadic polycystic kidney disease from 27 multicenter sites who lacked a positive family history for PKD. A total of 118 individuals were subjected to gene panel screening, encompassing 69 genes (version 1) or 92 genes (version 2) associated with the genetic causes of cystic kidney diseases (Figure 1). The detailed panel screening results of this cohort were reported in Fujimaru T et al. ^13, 26^. The forty-nine cases (42%) that remained unresolved after primary screening with targeted NGS panels were divided into three categories based on the type of variants detected. Briefly, the group in which only a mono-allelic variant of possible pathological significance in recessive genetic disease was detected was subdivided into Category 1 (previously reported pathological mutations) and Category 2 (pathogenic or likely-pathogenic based on ACMG criteria^14^). The group with no candidate variants or likely-benign variants was categorized as Category 3. The clinical information on the 49 cases unresolved in the primary screening is presented in Supplementary table S3. For Categories 1 and 2, targeted long-read sequencing (T-LRS) on the Oxford Nanopore platform was employed to identify variants in the contralateral allele^27^. In two cases (PT1049 and PT1192), an initially undetected deletion variant in *NPHP1* was identified, ultimately contributing to the diagnosis (Supplementary table S4). Whole-genome sequencing (WGS) was performed on 47 cases, including 8 that remained unresolved after T-LRS analysis and 39 Category 3 cases. T-LRS analysis and WGS were performed in collaboration with the University of Washington Center for Rare Disease Research. (UW-CRDR). Details regarding the analysis procedures, cutoff values used in various filtering steps, and other specifics are described in the Materials and Methods section and Supplementary Information. The details of the pathogenic variants detected are presented in Supplementary table S3, including two cases with *PKD1* mutations and one with a *PKD2* mutation, both of which were deletion variants. While the three *IFT140* mutations had been identified during the primary panel screening phase, there was no consensus at that time that mono-allelic variants in *IFT140* were a cause of PKD^6^. These three cases were redefined as having PKD due to the pathogenic variant in *IFT140*.

**Figure 1.**
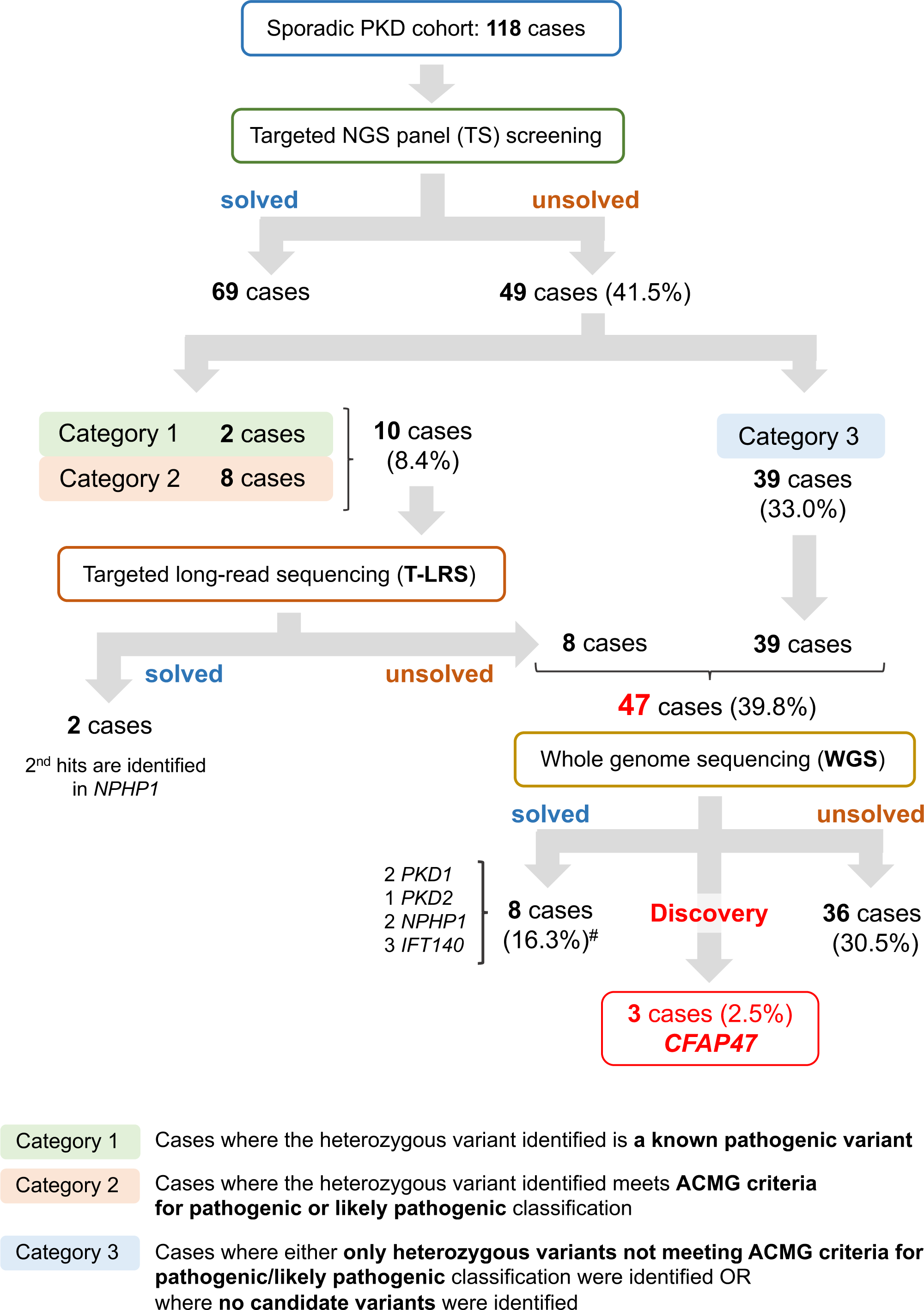
Summary of genetic analysis for a cohort of 118 cases with sporadic PKD. Targeted NGS panel analysis covering cystic kidney disease-related genes was followed by targeted long-read sequencing (T-LRS) and whole-genome sequencing (WGS). This approach identified *CFAP47* mutations in three cases, constituting 2.5% of the total cohort. ^#^The proportion within the 49 unsolved cases.

In addition to evaluating known genes, we sought to discover novel responsible genes for PKD using WGS. As the cohort involved simplex cases, candidate variants were filtered assuming either autosomal recessive (homozygous or compound heterozygous variants) or X-chromosome inheritance. This identified a rare hemizygous missense variant in *CFAP47*, an X-linked gene, in three male subjects (Figure 1, 2). The clinical course of these three patients is outlined below, and key clinical findings are summarized in Table 1. The pedigree and abdominal MRI images are presented in Figure 2.

**Figure 2.**
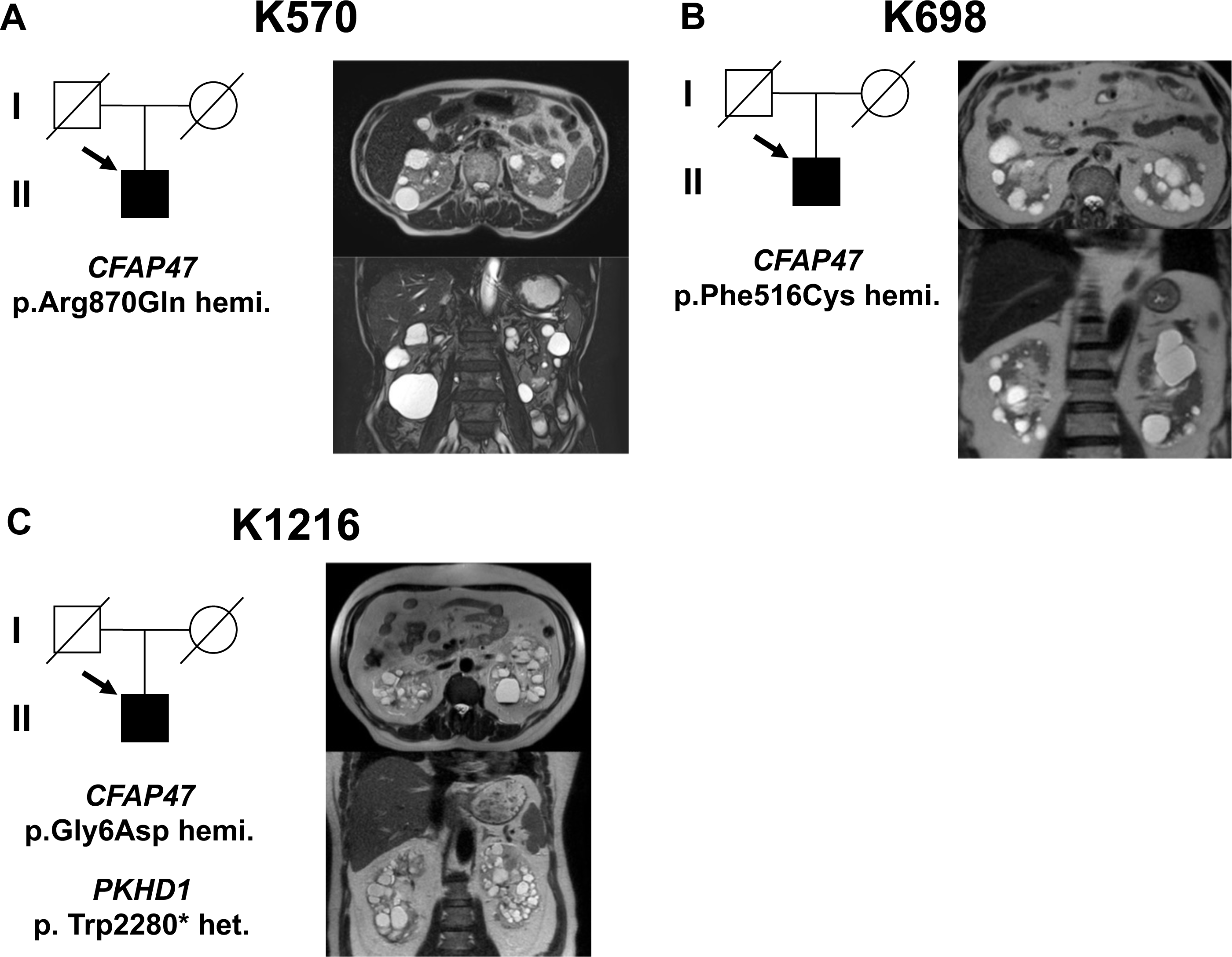
Family tree and MRI of Polycystic Kidneys. Family trees, genetic mutations, and abdominal MRI images of K570 (Case 1), K698 (Case 2), and K1216 (Case 3) are shown in Panels A, B, and C, respectively.

**Table 1.** Clinical characteristics of the three patients.

| Patient ID | Age<br>(range),<br>year | Sex | Hypertension | PLD | eGFR, ml/min/1.73m<sup>2</sup> | TKV, ml |
| --- | --- | --- | --- | --- | --- | --- |
| PT570 (Case 1) | 71-74 | Male | + | – | 53.5 | 587 |
| PT698 (Case 2) | 80-84 | Male | + | – | 21.8 | 698 |
| PT1216 (Case 3) | 55-60 | Male | + | – | 20.2 | 720 |

PLD, Polycystic liver disease.

**Table 2.** Profile of the candidate variants in the three.

| Patient ID. | Gene<sup>a</sup> | Variant | Zygosity | gnomAD<sup>b</sup> | CADD<sup>c</sup> | REVEL<sup>d</sup> | ACMG criteria | Reports |
| --- | --- | --- | --- | --- | --- | --- | --- | --- |
| PT570 | *CFAP47* | c.2609G>A; p.(Arg870Gln) | hemi. | 0.0003823 | 19.2 | 0.106 | VUS | none |
| PT698 | *CFAP47* | c.1547T>G; p.(Phe516Cys) | hemi. | 0.000006981 | 23.8 | 0.346 | VUS | none |
| PT1216 | *CFAP47* | c.17G>A; p.(Gly6Asp) | hemi. | 0.0003047 | 18.9 | 0.025 | VUS | none |
|  | *PKHD1* | c.6840G>A : p.(Trp2280*) | het. | N/A | 37.0 | N/A | Pathogenic | Hou L., et al. 2018<sup>32</sup> |

<sup>a</sup> NCBI accession in *CFAP47* and *PKHD1* are NM\_001304548.2 and NM\_138694.4, respectively.
<sup>b</sup> Genome Aggregation Database, v4.0 or v2.1.1<sup>24</sup>
<sup>c</sup> Combined Annotation-Dependent Depletion phred score<sup>18</sup>.
<sup>d</sup> REVEL score<sup>42</sup>
N/A, not available; hemi., hemizygous; het., heterozygous; VUS, variant of unknown significance

### Case presentation

The mean age at diagnosis for the three patients was approximately 70s, and renal function averaged 31.8 ± 18.8 ml/min/1.73 m² in terms of estimated Glomerular Filtration Rate (eGFR). The mean volume of both kidneys was 668.3 ± 71.3 mL, only mildly enlarged compared to the normal volume of 404 mL in males^28^. All patients presented with no liver cysts, while all were undergoing treatment for hypertension. A brief summary of each case is provided below.

#### Case 1 (PT570)

A man in his 70s, under treatment with Angiotensin II Receptor Blockers (ARBs) for hypertension, maintained a regular blood pressure around 130/80 mmHg. He presented with bilateral renal cysts and mild proteinuria (urine total protein to creatinine ratio, UPCR: ∼0.8g/gCr). Throughout the disease course, urine occult blood remained consistently negative. Magnetic Resonance Angiography (MRA) of the head revealed no cerebral aneurysm, and hepatic cysts were absent (Figure 2A). Renal function had remained stable for several years, with serum creatinine ranging from 1.05 to 1.15 mg/dl. Although one of the parents passed away due to cerebral hemorrhage, no renal disease was detected. Furthermore, the any children showed no signs of renal cysts.

#### Case 2 (PT698)

An man in his 80s a history of urinary tract stone disease, benign prostatic hyperplasia, and aortic stenosis. He was prescribed ARBs and Calcium Channel Blockers (CCB) for hypertension, maintaining a blood pressure of 125/75 mmHg. There was no family history of renal disease, including cystic kidney disease. His family doctor reported renal dysfunction, noting a decreased estimated glomerular filtration rate (eGFR) of 50.4 ml/min/1.73 m² and UPCR of 0.5 g/gCr. His renal function deteriorated further to Blood Urea Nitrogen (BUN) 28 mg/dl, Creatinine (Cr) 2.31 mg/dl, eGFR 22.1 ml/min/1.73 m², and UPCR 1.5 g/gCr. UPCR increased to about 1 g/gCr, but urinary occult blood remained negative. Abdominal MRI revealed multiple cysts in both kidneys, with no hepatic cysts (Figure 2B). Echocardiography indicated no left ventricular contractility issues, only mild aortic stenosis (AS).

#### Case 3 (PT1216)

A man in his late 50s with hypertension, prescribed ARB and CCB to maintain a stable blood pressure around 130/80 mmHg. He is also taking warfarin for persistent atrial fibrillation and has a history of bilateral kidney stones. Urine analysis revealed mild proteinuria with a UPCR of approximately 0.6 g/gCr and mild urine occult blood (2+ in qualitative analysis). Abdominal MRI displayed bilateral multiple renal cysts but no liver cysts (Figure 2C). One of his parent passed away from lung cancer, with no renal cysts noted. The other parent did not have any documented renal disease. His older sibling passed away shortly after birth, and the cause remains unknown. No renal cysts were observed in his children. Echocardiography revealed no left ventricular contractility problems and no apparent valvular disease.

### Interpretation of the Pathogenicity of *CFAP47* Variants

*CFAP47* encodes Cilia And Flagella Associated Protein 47, a protein that plays a role in the formation and function of cilia and flagella ^25, 29^. Given the association between genes underlying cystic kidney disease and genes encoding cilia-related proteins^30^, we considered *CFAP47* as a high-priority candidate gene.

We identified three rare missense variants in *CFAP47* (NM_001304548.2): c.2609G>A; p.(Arg870Gln), c.1547T>G; p.(Phe516Cys), c.17G>A; p.(Gly6Asp) in Case 1 to 3, respectively (Table 3). The affected individuals were all males with hemizygous X-chromosome mutations, exhibiting an allele frequency of less than 0.0005 in gnomAD. *In silico* pathogenicity prediction scores, CADD and REVEL, generally favored the damaging variant.

Application of ACMG criteria^14^ classified all three variants as VUS (variants of unknown significance), and there was no record of any of these variants in ClinVar^31^. In Case 3 (PT1216), a heterozygous nonsense mutation (*PKHD1* (NM_138694.4): c.6840G>A: p.(Trp2280*))^32^, previously reported as a disease-causing mutation in *PKHD1*, the gene associated with ARPKD, was identified. We used T-LRS to exclude a second pathogenic variant in the contralateral allele. This variant is also interpreted as pathogenic (PVS1, PM2, PP3, PP5) by ACMG criteria and is considered to have pathological significance. However, given that it is a monoallelic variant and ARPKD is an autosomal recessive disease, the extent to which the phenotype is affected, if at all, is unclear.

### Basic Examination of the Effects of CFAP47 on the Renal Phenotype

Considering the numerous uncertainties in the molecular biology of CFAP47, including its renal localization, and the interpretation of VUS on a novel gene, we chose to conduct a basic experimental validation to confirm whether *CFAP47* is indeed the causative gene. Our investigation confirmed the expression of CFAP47 in human renal tubular cilia through double staining with antibodies to CFAP47 and acetylated alpha-tubulin, a marker of primary cilia (Figure 2A). Incidentally, immunohistochemistry for CFAP47 was also attempted in mouse kidneys; however, despite using multiple antibodies and employing various techniques, it proved unattainable. Subsequently, we examined the renal phenotype of *Cfap47* knock-out (KO) mice (*Cfap47^-/Y^*). This mouse model, previously reported by Chunyu Liu and Feng Zhang et al. as a pathological model of human asthenoteratozoospermia^25^, served as the basis for our collaborative study analysis. As illustrated in Figure 3B and 3C, the kidneys of 40-week-old KO mice were slightly but significantly larger than those of wild-type (WT) (*Cfap47^+/Y^*) mice, although no gross macro-morphological cyst formation was evident. Histological examination using HE staining revealed scattered areas with sparing HE staining in the kidneys of the KO mice, particularly in the subcortical and corticomedullary borders (3D, left). Similar changes were consistently observed in all three individual KO mice. At higher magnification, vacuolation of tubular cells was evident in KO mice, along with the fusion of some vacuoles, displaying tubular dilatation-like features (3D, right). Vacuolation of tubular epithelial cells was not observed in WT mice. The vacuolation and tubular dilatation were considered to contribute to the mild enlargement of the kidney volume. This suggests that the loss of CFAP47 function induces cyst-like changes in renal tubules.

**Figure 3.**
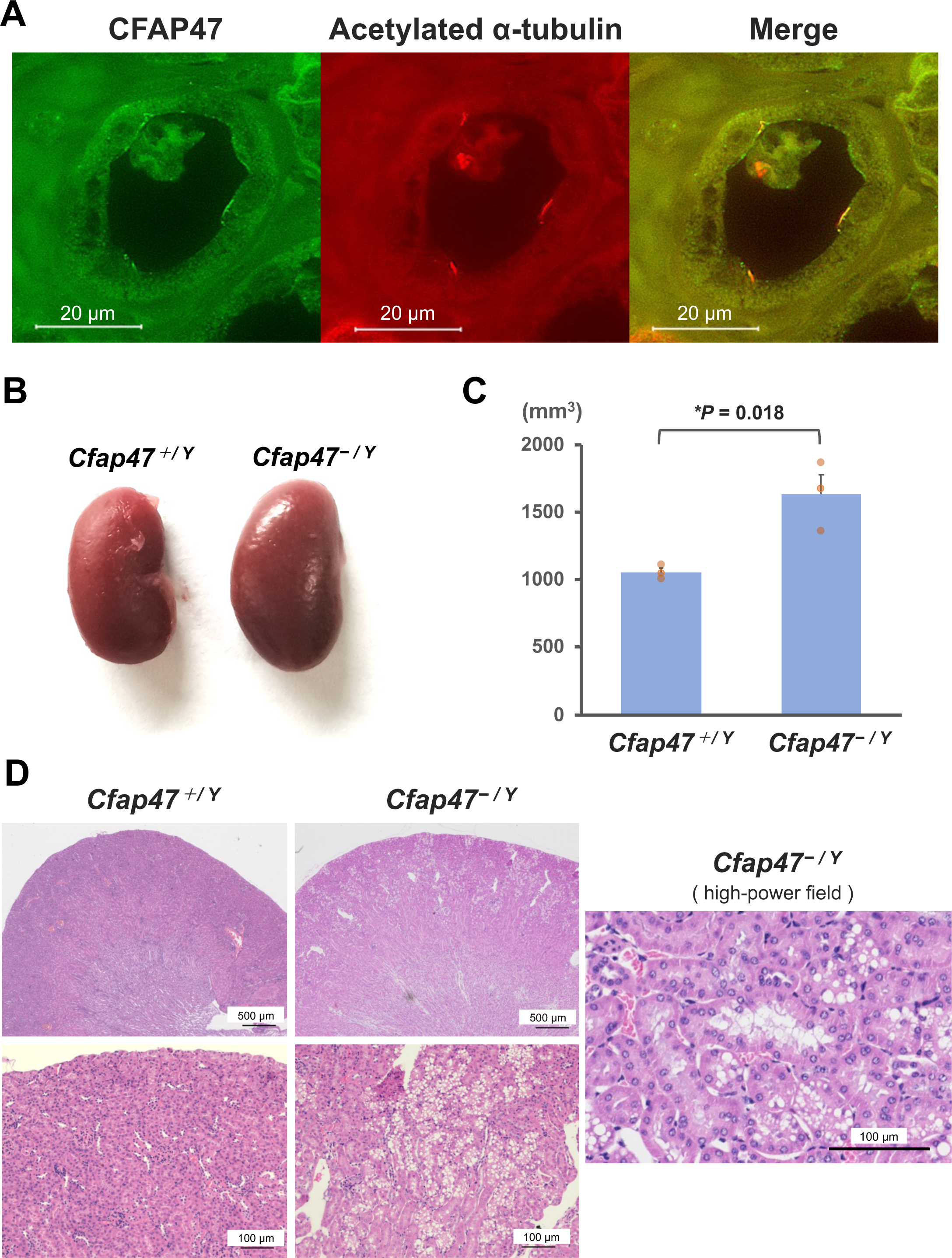
Evaluation of CFAP47 expression sites in human kidney tubules and renal morphology of *Cfap47^-/Y^* mice. (A) Human kidney normal tubules co-immunostained with acetylated alpha-tubulin antibody (red) and CFAP47 antibody (green); arrowheads indicate positive staining sites; (B) *Cfap47^+/Y^* and *Cfap47^-/Y^* mouse kidney macro-morphology images; (C) Kidney Volume Comparison using the ellipsoid formula: (length × width × [π/6])^2^ (D) Low and high magnification images with Hematoxylin-Eosin Stain.

## Discussion

A comprehensive panel screening of causative genes for cystic kidney disease was conducted on a cohort of 118 patients with sporadic cases of cystic kidney disease and no familial history. WGS was performed on 47 patients who did not exhibit an identified responsible variant. Rare hemizygous missense variants in *CFAP47* were identified in three men within this subgroup. Basic verification, guided by histological evaluation, confirmed the localization of CFAP47 to the primary cilia of human renal tubular epithelial cells. Notably, tubular epithelial cells in *Cfap47*-KO mice exhibited evidence of vacuolation and tubular dilation. These findings suggest that *CFAP47* serves as a causative gene for X-linked cystic kidney disease.

The genetic background analysis of sporadic PKD in our cohort has been previously reported^13, 26^. As presented in Figure 1, among the unsolved cases, the 47 individuals constituted 39.8% of the total 118 cases, and the three individuals carrying the *CFAP47* pathogenic variants accounted for approximately 2.5% of the entire cohort. As a result, diagnostic variants were identified in 71 cases (61%) in the cohort.

X-linked cystic kidney disease has previously been associated with oral-facial-digital syndrome type 1 (*OFD1*), characterized by malformations in the face, oral cavity, and digits. Additionally, the clinical phenotype often involves mental retardation and renal functional impairment^33^. In contrast, the three patients with *CFAP47* mutations in this study exhibited no facial deformities or mental retardation and lacked extra-renal manifestations. The patient is elderly, and since both parents have already passed away, it is challenging to conduct familial analysis. However, if the mutation is not *de novo*, one of the parents of the proband is likely to be an obligate carrier of the same mutation. In females with X-linked inheritance, the pathogenic impact of the gene mutation is further influenced by X chromosome inactivation, leading to diverse phenotypic expressions. In the three cases of this study, the parents did not exhibit clear symptoms of renal cysts or renal failure, which is consistent with the variability in the manifestation of X-linked inheritances.

The pathological significance interpretation of the detected mutations based on the ACMG criteria was all classified as VUS. Additionally, due to the incomplete supporting evidence from *in silico* pathogenicity prediction scores, observations of the kidneys in *Cfap47*-KO mice were conducted. There is ongoing discussion regarding the interpretation of VUS^34^, especially in cases of low-penetrance and recessive disease-associated variants, or when the associated phenotypes only partially overlap with the originally reported disease phenotypes^35^. In fact, reconsideration attempts of VUS classification have shown that some were not interpretable as benign^36^. The variants identified in Cases 1 and 3 do not necessarily exhibit ultra-rare MAFs in gnomAD (0.00038 and 0.00030, respectively). However, given the mild expression phenotype, it is possible that these variants may not have been recognized as chronic kidney disease (CKD) or PKD until old age.

The relationship between vacuolation of tubular epithelial cells and the development of large cysts remains unclear, but vacuolation itself has been reported to occur due to various factors such as ischemia, hyperosmolarity, and lipid accumulation^37–39^. Although this study cannot completely negate some of the aforementioned background factors, the absence of any tissue changes in the littermate wild-type mice (*Cfap47^+/Y^*) suggests that the observed alterations are associated with the loss of CFAP47 function. In general, mouse models of cystic kidney disease, even with the same genetic background, exhibit phenotypes that are milder and progress more slowly than the actual phenotypes observed in humans. This is particularly true for recessive forms of the model, adding further complexity to the interpretation^40^. Considering the presence of partially fused vacuoles and areas displaying dilatation of tubular lumens, it is conceivable that vacuolation may contribute to the initiation of cyst formation. It should be noted that the CFAP47 antibody used in this study was not suitable for staining mouse kidneys due to sensitivity issues, making detailed histological examinations in mouse kidneys challenging.

In the context of considering the mechanism underlying cyst formation due to *CFAP47* mutations, previously reported minor causative gene groups of ADPKD and most ADPLD proteins are involved in the folding and transport of proteins in the endoplasmic reticulum (ER). Among these proteins, PC-1 (Polycystin-1) has been identified as particularly sensitive to the reduction in the dosage of these proteins causative for cystic diseases^5, 8, 41^. For example, *GANAB*, a causative gene for ADPKD, encodes the glucosidase II subunit α (GIIα), which is necessary for the maturation, surface expression, and ciliary localization of ADPKD proteins, PC-1 and PC-2^5^. Similarly, *CFAP47* mutations may also result in negative effects on PC-1/PC-2, but further investigation is necessary.

The cysts in PT1216 exhibited some morphological differences compared to the other two individuals, who displayed numerous small cysts, and more impaired renal function. In this case, along with the *CFAP47* mutation, a previously reported mutation in *PKHD1*, responsible for autosomal recessive polycystic kidney disease (ARPKD), was identified. The coexistence with *CFAP47* mutations may have had some impact on the more severe phenotype in this case.

Cases 1 and 3 had children, suggesting that their reproductive abilities were unaffected. Missense mutations reported as causes of asthenoteratozoospermia, namely S1742G, I2385N, and P2890T, are localized at the C-terminal end^25^. The difference in the localization of mutations may be associated with differences in phenotype.

The limitation lies in the selection of sporadic cases, as mentioned earlier, it is primarily based on medical history rather than necessarily on the imaging evaluation of all parents. This approach may include cases that do not strictly qualify as true sporadic cases. Although the KO mice provide an optimal model for studying the loss of function in CFAP47, it is important to note that they do not directly replicate the pathological effects associated with the specific missense mutations investigated in this study.

In conclusion, *CFAP47* is a newly discovered gene associated with PKD, demonstrating X-linked inheritance, a trait uncommon in cases of PKD. *CFAP47* should be considered as one of the causes of sporadic cases of PKD. Further accumulation of cases is warranted for a more comprehensive understanding.

## Acknowledgments

We would like to express our gratitude to all patients who participated in our study. Additionally, we extend our thanks to the doctors who provided us with the patient’s samples and corresponding clinical data included in this study. Sequencing and data analysis were provided by the University of Washington Center for Rare Disease Research (UW-CRDR). The content is solely the responsibility of the authors and does not necessarily represent the official views of the National Institutes of Health.

## Authors’ Contributions

Conceptualization: T.Mori, TF, SU, ES. Data curation: T.Mori, TF, KP, DEM, MZ, UW-CRDR. Formal analysis: T.Mori, TF, KP, DEM, MZ, UW-CRDR, ES. Funding acquisition: T.Mori, TF, SU, ES. Investigation: T.Mori, TF, CL, KP, KY, TS, MC, DEM, MZ, UW-CRDR, FZ, ES. Methodology: T.Mori, TF, SU, ES. Resources: T.Mori, TF, CL, KP, KY, TS, MC, SM, FA, YM, HK, KS, UW-CRDR, JXC, MJB, YQT, FZ, SU, ES. Supervision: TM, YQT, FZ, SU, ES. Writing-original draft: T.Mori, TF, ES. Writing-review & editing: T.Mori, TF, CL, KP, KY, TS, MC, DEM, MZ, SM, FA, YM, HK, KS, UW-CRDR, JXC, MJB, YQT, FZ, SU, ES.

## Data Availability Statement

The data underlying this article cannot be shared publicly due to the privacy of individuals that participated in the study. The data will be shared on reasonable request to the corresponding author.

## References

1. Rossetti S, Consugar MB, Chapman AB, et al. Comprehensive molecular diagnostics in autosomal dominant polycystic kidney disease. J Am Soc Nephrol 2007; 18: 2143–2160.

2. Mallawaarachchi AC, Lundie B, Hort Y, et al. Genomic diagnostics in polycystic kidney disease: an assessment of real-world use of whole-genome sequencing. Eur J Hum Genet 2021; 29: 760–770.

3. Chang AR, Moore BS, Luo JZ, et al. Exome Sequencing of a Clinical Population for Autosomal Dominant Polycystic Kidney Disease. JAMA 2022; 328: 2412–2421.

4. Yu CC, Lee AF, Kohl S, et al. PKD2 founder mutation is the most common mutation of polycystic kidney disease in Taiwan. NPJ Genom Med 2022; 7: 40.

5. Porath B, Gainullin VG, Cornec-Le Gall E, et al. Mutations in GANAB, Encoding the Glucosidase IIα Subunit, Cause Autosomal-Dominant Polycystic Kidney and Liver Disease. Am J Hum Genet 2016; 98: 1193–1207.

6. Senum SR, Li YSM, Benson KA, et al. Monoallelic IFT140 pathogenic variants are an important cause of the autosomal dominant polycystic kidney-spectrum phenotype. Am J Hum Genet 2022; 109: 136–156.

7. Lemoine H, Raud L, Foulquier F, et al. Monoallelic pathogenic ALG5 variants cause atypical polycystic kidney disease and interstitial fibrosis. Am J Hum Genet 2022; 109: 1484–1499.

8. Cornec-Le Gall E, Olson RJ, Besse W, et al. Monoallelic Mutations to DNAJB11 Cause Atypical Autosomal-Dominant Polycystic Kidney Disease. Am J Hum Genet 2018; 102: 832–844.

9. Besse W, Chang AR, Luo JZ, et al. Mutation Carriers Develop Kidney and Liver Cysts. J Am Soc Nephrol 2019; 30: 2091–2102.

10. Matsuo S, Imai E, Horio M, et al. Revised equations for estimated GFR from serum creatinine in Japan. Am J Kidney Dis 2009; 53: 982–992.

11. Cadnapaphornchai MA, McFann K, Strain JD, et al. Prospective change in renal volume and function in children with ADPKD. Clin J Am Soc Nephrol 2009; 4: 820–829.

12. Mori T, Hosomichi K, Chiga M, et al. Comprehensive genetic testing approach for major inherited kidney diseases, using next-generation sequencing with a custom panel. Clin Exp Nephrol 2017; 21: 63–75.

13. Fujimaru T, Mori T, Sekine A, et al. Kidney enlargement and multiple liver cyst formation implicate mutations in PKD1/2 in adult sporadic polycystic kidney disease. Clin Genet 2018; 94: 125–131.

14. Richards S, Aziz N, Bale S, et al. Standards and guidelines for the interpretation of sequence variants: a joint consensus recommendation of the American College of Medical Genetics and Genomics and the Association for Molecular Pathology. Genet Med 2015; 17: 405–424.

15. Li J, Lupat R, Amarasinghe KC, et al. CONTRA: copy number analysis for targeted resequencing. Bioinformatics 2012; 28: 1307–1313.

16. Li H. Minimap2: pairwise alignment for nucleotide sequences. Bioinformatics 2018; 34: 3094–3100.

17. McLaren W, Gil L, Hunt SE, et al. The Ensembl Variant Effect Predictor. Genome Biol 2016; 17: 122.

18. Kircher M, Witten DM, Jain P, et al. A general framework for estimating the relative pathogenicity of human genetic variants. Nat Genet 2014; 46: 310–315.

19. de Sainte Agathe JM, Filser M, Isidor B, et al. SpliceAI-visual: a free online tool to improve SpliceAI splicing variant interpretation. Hum Genomics 2023; 17: 7.

20. Sedlazeck FJ, Rescheneder P, Smolka M, et al. Accurate detection of complex structural variations using single-molecule sequencing. Nat Methods 2018; 15: 461–468.

21. Heller D, Vingron M. SVIM: structural variant identification using mapped long reads. Bioinformatics 2019; 35: 2907–2915.

22. Van der Auwera GA, Carneiro MO, Hartl C, et al. From FastQ data to high confidence variant calls: the Genome Analysis Toolkit best practices pipeline. Curr Protoc Bioinformatics 2013; **43**: 11.10.11–11.10.33.

23. Pais LS, Snow H, Weisburd B, et al. seqr: A web-based analysis and collaboration tool for rare disease genomics. Hum Mutat 2022; 43: 698–707.

24. Chen S, Francioli LC, Goodrich JK, et al. A genomic mutational constraint map using variation in 76,156 human genomes. Nature 2024; 625: 92–100.

25. Liu C, Tu C, Wang L, et al. Deleterious variants in X-linked CFAP47 induce asthenoteratozoospermia and primary male infertility. Am J Hum Genet 2021; 108: 309–323.

26. Fujimaru T, Kawanishi K, Mori T, et al. Genetic Background and Clinicopathologic Features of Adult-onset Nephronophthisis. Kidney Int Rep 2021; 6: 1346–1354.

27. Miller DE, Sulovari A, Wang T, et al. Targeted long-read sequencing identifies missing disease-causing variation. Am J Hum Genet 2021; 108: 1436–1449.

28. Cheong B, Muthupillai R, Rubin MF, et al. Normal values for renal length and volume as measured by magnetic resonance imaging. Clin J Am Soc Nephrol 2007; 2: 38–45.

29. Liao HQ, Guo ZY, Huang LH, et al. WDR87 interacts with CFAP47 protein in the middle piece of spermatozoa flagella to participate in sperm tail assembly. Mol Hum Reprod 2022; 29.

30. Ma M. Cilia and polycystic kidney disease. Semin Cell Dev Biol 2021; 110: 139–148.

31. Landrum MJ, Lee JM, Riley GR, et al. ClinVar: public archive of relationships among sequence variation and human phenotype. Nucleic Acids Res 2014; 42: D980–985.

32. Hou L, Du Y, Zhang M, et al. Novel mutations of. Int J Clin Exp Pathol 2018; 11: 2869–2874.

33. Feather SA, Woolf AS, Donnai D, et al. The oral-facial-digital syndrome type 1 (OFD1), a cause of polycystic kidney disease and associated malformations, maps to Xp22.2-Xp22.3. Hum Mol Genet 1997; 6: 1163–1167.

34. Aklilu AM, Gulati A, Kolber KJ, et al. The VUS Challenge in Cystic Kidney Disease: A Case-Based Review. Kidney360 2024; 5: 152–159.

35. Houge G, Laner A, Cirak S, et al. Stepwise ABC system for classification of any type of genetic variant. Eur J Hum Genet 2022; 30: 150–159.

36. Tsai GJ, Rañola JMO, Smith C, et al. Outcomes of 92 patient-driven family studies for reclassification of variants of uncertain significance. Genet Med 2019; 21: 1435–1442.

37. Zhou C, Yool AJ, Nolan J, et al. Armanni-Ebstein lesions: a need for clarification. J Forensic Sci 2013; 58 **Suppl 1**: S94–98.

38. Zhou C, Vink R, Byard RW. Hyperosmolarity Induces Armanni-Ebstein-like Renal Tubular Epithelial Swelling and Cytoplasmic Vacuolization. J Forensic Sci 2017; 62: 229–232.

39. Parai JL, Kodikara S, Milroy CM, et al. Alcoholism and the Armanni-Ebstein lesion. Forensic Sci Med Pathol 2012; 8: 19–22.

40. Sieben CJ, Harris PC. Experimental Models of Polycystic Kidney Disease: Applications and Therapeutic Testing. Kidney 360 2023; 4: 1155–1173.

41. Fedeles SV, Tian X, Gallagher AR, et al. A genetic interaction network of five genes for human polycystic kidney and liver diseases defines polycystin-1 as the central determinant of cyst formation. Nat Genet 2011; 43: 639–647.

42. Ioannidis NM, Rothstein JH, Pejaver V, et al. REVEL: An Ensemble Method for Predicting the Pathogenicity of Rare Missense Variants. Am J Hum Genet 2016; 99: 877–885.

