## Supplementary Information for "*CFAP47* is a novel causative gene implicated in X-linked polycystic kidney disease"

1 Department of Nephrology, Graduate School of Medical and Dental Sciences, Tokyo Medical and Dental University, Tokyo, Japan.
2 Soong Ching Ling Institute of Maternal and Child Health, International Peace Maternity and Child Health Hospital, Shanghai Jiao Tong University School of Medicine, Shanghai, China.
3 State Key Laboratory of Genetic Engineering,, Institute of Medical Genetics and Genomics, Fudan University, Shanghai, China.
4 Department of Genome Sciences, University of Washington, Seattle, WA, USA.
5 Department of Comprehensive Pathology, Graduate School of Medical and Dental Sciences, Tokyo Medical and Dental University, Tokyo, Japan.
6 Department of Nephrology and Rheumatology, Toranomon Hospital, Japan.
7 Okinaka Memorial Institute for Medical Research, Toranomon Hospital, Tokyo, Japan.
8 Department of Pediatrics, Division of Genetic Medicine, University of Washington, 1959 NE Pacific Street, Box 357371, Seattle, WA, 98195, USA.
9 Brotman-Baty Institute for Precision Medicine, 1959 NE Pacific Street, Box 357657, Seattle, WA, 98195, USA.
10 Institute of Reproductive and Stem Cell Engineering, NHC Key Laboratory of Human Stem Cell and Reproductive Engineering, School of Basic Medical Science, Central South University, Changsha, China.
11 Clinical Research Center for Reproduction and Genetics in Hunan Province, Reproductive and Genetic Hospital of CITIC-Xiangya, Changsha, China.
*contributed equally

Corresponding author: Eisei Sohara
Department of Nephrology, Graduate School of Medical and Dental Sciences
Tokyo Medical and Dental University
1-5-45 Yushima, Bunkyo, Tokyo 113-8519, Japan


**University of Washington Center for Rare Disease Research (UW-CRDR)**
Chia-Lin Wei[1], Michael J. Bamshad[1,2] and Evan E. Eichler[1,3]

Kailyn Anderson[1], Peter Anderson[1], Tamara J. Bacus[1], Sabrina Best[1], Elizabeth E. Blue[1], Katherine Brower[1], Kati J. Buckingham[1], Brianne Carroll[1], Silvia Casadei[1], Jessica X. Chong[1], Nikhita Damaraju[1], Colleen P. Davis[1], Christian D. Frazar[1], Sophia Gibson[1], Joy Goffena[1], William W. Gordon[1], Jonas A. Gustafson[1], William T. Harvey[1], Martha Horike-Pyne[1], Jameson R. Hurless[1], Caitlin Jacques[1], Gail P. Jarvik[1], Eric Johanson[1], J. Thomas Kolar[1], Xiaomeng Liu[1], Colby T. Marvin[1], Sean McGee[1], Holli Meyers[1], Heather Mefford[4], Danny E. Miller[1,2], Patrick M. Nielsen[1], Karynne Patterson[1], Aparna Radhakrishnan[1], Matthew A. Richardson[1], Erica L. Ryke[1], Aliya Sarkytbayeva[1], Tristan Shaffer[1], Kathryn M. Shively[1], Olivia M. Sommers[1], Sophie H.R. Storz[1], Joshua D. Smith[1], Lea M. Starita[1], Monica Tackett[1], Sydney A. Ward[1], Jeffrey M. Weiss[1], Qian Yi[1], and Miranda P.G. Zalusky[1].

1 University of Washington
2 Seattle Children’s Hospital
3 Howard Hughes Medical Institute
4 St. Jude Children’s Research Hospital

***Genetic analysis***

**Purification of Genomic DNA from Blood Samples**

Genomic DNA in blood leukocytes was purified using the QIAamp DNA Blood Mini Kit (Qiagen, Hilden, Germany). A quality check was conducted with a spectrophotometer DU700 (Beckman Coulter, Brea, California) to ensure that the A260/A280 ratio met or exceeded 1.8. Concentration measurements were performed using a Qubit Fluorometer (Thermo Fisher Scientific, Waltham, Massachusetts).

**Capture-based sequencing**

Biotinylated RNA capture probes for the coding regions of the 69 (panel version 1) or 92 (panel version 2) genes associated with inherited renal cystic diseases such as ADPKD, autosomal recessive polycystic kidney disease (ARPKD), nephronophthisis-related ciliopathy (including Joubert syndrome, Meckel syndrome, Senior-Løken syndrome, Bardet-Biedl syndrome, and skeletal ciliopathy), autosomal dominant tubulointerstitial kidney disease, autosomal dominant polycystic liver disease (ADPLD), and other renal cystic diseases (Supplementary Table S1) were designed using Agilent's SureDesign service (https://earray.chem.agilent.com/suredesign/home.htm 27th Jan, 2024 last accessed). More details can be found in our previous reports[1-3]. Pooled and barcoded libraries for next-generation sequencing were prepared using SureSelect QXT kits (Agilent Technologies, Santa Clara, California), following the manufacturer's protocol. The prepared libraries were sequenced with 150-bp single-end reads using MiSeq sequencers (Illumina, San Diego, California). The original read files were aligned to a human reference sequence (hg19hs37d5) using BWA-MEM (Burrows-Wheeler Aligner; v0.7.15) [4], and SAM files were generated. Subsequently, the files were sorted and indexed using SAMtools (v.0.1.19) [5]. All aligned read data are subject to the following steps: (1) "duplicate removal" is performed, (i.e. the removal of reads with duplicate start positions; Picard MarkDuplicates; v0.1.19) and (2) base qualities are recalibrated (GATK BaseRecalibrator; v3.3) [6]. Variant detection and genotyping are performed using the HaplotypeCaller (HC) tool from GATK (v3.3).

Candidate variant filtering was conducted as previously described[7]. Initially, we excluded single nucleotide variants (SNVs) with allele frequencies exceeding 0.01 in any population within the gnomAD database (v2.1.1) [8]; National Heart, Lung, and Blood Institute Exome Sequencing Project exome variant server dataset ESP6500 (http://evs.gs.washington.edu/EVS/ 31st Aug 2020 last accessed); or the allele frequency panel of 38,000 Japanese individuals from The Tohoku Medical Megabank Organization (https://jmorp.megabank.tohoku.ac.jp, 27th Jan, 2024 last accessed). We excluded single nucleotide variants (SNVs) with "LOW" impact severities based on the definition in the GEMINI software[9]. This category encompassed various functional predictions, including "synonymous_coding," "intergenic," "upstream," "UTR," "intron," etc. For interpreting the significance of the variants, we employed CADD[10], SIFT[11], Polyphen-2[12], MCAP[13], and GERP++ conservation scores[14]. To assess the current evidence on the pathogenicity of previously reported variants, we reviewed the literature available in the Human Gene Mutation Database Pro (https://www.hgmd.cf.ac.uk/ac/index.php 27th Jan, 2024 last accessed).

**Whole genome sequencing analysis**

Library Production

Starting with a minimum of 750ng of DNA, samples are sheared in a 96-well format using a LE220 (Covaris, Woburn, Massachusetts) focused ultrasonicator targeting 380 bp inserts. The resulting sheared DNA is cleaned with Agencourt AMPure XP beads (Beckman Coulter, Brea, California) to remove sample impurities prior to library construction. Shearing is followed by size selection and sample prep is performed using the KAPA Hyper Prep kit (KR0961 v1.14, KAPA Bio systems, Wilmington, Massachusetts). End-repair, A-tailing, and ligation are performed as directed. Two final AMPure cleanups are performed after ligation in order to remove excess adapter dimers from the library. All library construction steps are automated on the Janus platform (Perkin Elmer, Waltham,

Massachusetts). Library yield is quantified using Quant-IT dsDNA High Sensitivity kit (Q33120, Invitrogen, Waltham, Massachusetts). Libraries are validated in triplicate using the CFX384 Real-Time System (Bio-Rad Laboratories, Hercules, California) and KAPA Library Quantification Kit (KK4824, KAPA Bio systems, Wilmington, Massachusetts).

Clustering/Sequencing

Barcoded genome libraries are pooled using liquid handling robotics prior to loading. Massively parallel sequencing-by-synthesis with fluorescently labeled, reversibly terminating nucleotides is carried out on the NovaSeq sequencer (Illumina, San Diego, California).

Read Processing

Processing pipeline consists of the following elements: (1) base calls generated in real-time on the NovaSeq6000 instrument (RTA 3.1.5, Illumina, San Diego, California); (2) demultiplexed, unaligned BAM files produced by Picard ExtractIlluminaBarcodes and IlluminaBasecallsToSam; and (3) BAM files aligned to a human reference (hg19hs37d5) using BWA-MEM (Burrows-Wheeler Aligner; v0.7.15) [4, 5]. Read data from a flow-cell lane is treated independently for alignment and QC purposes in instances where the merging of data from multiple lanes is required (e.g., for sample multiplexing). All aligned read data are subject to the following steps: (1) "duplicate removal" is performed, (i.e. the removal of reads with duplicate start positions; Picard MarkDuplicates; v2.6.0) and (2) base qualities are recalibrated (GATK BaseRecalibrator; v3.7) [6].

Variant Detection

Variant detection and genotyping are performed using the HaplotypeCaller (HC) tool from GATK (3.7). Variant data for each sample are formatted (variant call format [VCF]) as "raw" calls that contain individual genotype data for one or multiple samples and flagged using the filtration walker (GATK) to mark sites that are of lower quality/false positives [e.g., low quality scores (Q50), allelic imbalance

(ABHet 0.75), long homopolymer runs (HRun> 4) and/or low quality by depth (QD < 5)].

Data Analysis QC

All sequence data undergo a QC protocol. For whole genomes, this includes an assessment of: (1) mean coverage; (2) fraction of genome covered greater than 10X; (3) duplicate rate; (4) mean insert size; (5) contamination ratio; (6) mean Q20 base coverage; (7) Transition/Transversion ratio (Ti/Tv); (8) fingerprint concordance > 99%; (9) sample homozygosity and heterozygosity; and (10) sample contamination < 2%. All QC metrics for both single-lane and merged data are reviewed by a sequence data analyst to identify data deviations from known or historical norms. Lanes/samples that fail QC are flagged in the system and can be re-queued for library prep (< 1% failure) or further sequencing (< 2% failure), depending upon the QC issue. Genome completion is defined as having > 95% of the target at > 10X coverage and > 90% of the target at > 20X coverage. Typically this requires mean coverage at 30X.

Variant Annotation

Automated pipeline for annotation of variants derived from genome data, the SeattleSeq Annotation Server (http://gvs.gs.washington.edu/SeattleSeqAnnotation/ 31st Aug, 2020 last accessed) was used. This publicly accessible server returns annotations including dbSNP rsID (or whether the coding variant is novel), gene names and accession numbers, predicted functional effect (e.g., splice-site, nonsynonymous, missense, etc.), protein positions and amino-acid changes, PolyPhen predictions, conservation scores (e.g., PhastCons, GERP), ancestral allele, dbSNP allele frequencies, and known clinical associations. The annotation process has also been automated into our analysis pipeline to produce a standardized, formatted output (variant call format [VCF]). Variant filtering was conducted using seqr, a web-based Whole Genome Sequencing (WGS) analysis tool developed by the Broad Institute[15].

**Supplementary table S1. Targeted genes and disease categories included in the panels**

| Disease | Genes |
|---|---|
| ADPKD | *PKD1, PKD2, GANAB*[a] |
| ARPKD | *PKHD1, DZIP1L*[a] |
| Nephronophthisis | *NPHP1, INVS, NPHP3, NPHP4, IQCB1, CEP290, GLIS2, RPGRIP1L, NEK8, SDCCAG8, TMEM67, TTC21B, WDR19, ZNF423, CEP164, ANKS6, IFT172, CEP83, DCDC2, XPNPEP3, SLC41A1, MAPKBP1*[a] |
| JBS | *NPHP1, CEP290, RPGRIP1L, TMEM67, TTC21B, ZNF423, CEP164, IFT172, INPP5E, TMEM216, AHI1, ARL13B, CC2D2A, OFD1, KIF7, TCTN1, TMEM237, CEP41, TMEM138, C5orf42, TCTN3, TMEM231, CSPP1, PDE6D, MKS1, TCTN2, B9D1 ARMC9*[a], *CEP104*[a], *CEP120*[a], *KIAA0556*[a], *KIAA0586*[a], *PIBF1*[a], *SUFU*[a], *TMEM107*[a] |
| MKS | *NPHP3, CEP290, RPGRIP1L, TMEM67, TMEM216, CC2D2A, TMEM231, MKS1, TCTN2, B9D1, B9D2, KIF14*[a], *TMEM107*[a] |
| SLS | *NPHP1, INVS, NPHP3, NPHP4, IQCB1, CEP290, GLIS2, SDCCAG8, WDR19, CEP164 TRAF3IP1*[a] |
| BBS | *CEP290, SDCCAG8, TMEM67, TTC21B, WDR19, IFT172, MKS1, BBS1, BBS2, ARL6, BBS4, BBS5, MKKS, BBS7, TTC8, BBS9, BBS10, TRIM32, BBS12, WDPCP, BBIP1, IFT27, CCDC28B, C8orf37*[a], *IFT74*[a] |
| Skeletal ciliopathy | *TTC21B, WDR19, IFT172, WDR35, IFT122, IFT140, IFT43* |
| ADTKD | *MUC1, UMOD, HNF1B, REN*[a], *SEC61A1*[a] |
| ADPLD | *PRKCSH*[a], *SEC63*[a], *ALG8*[a], *LRP5*[a], *SEC61B*[a], *GANAB*[a] |
| Others | *ASS1, NOTCH2, TSC2*[a] |

Panel version 1: 69 genes, and version 2: 92 genes ("a" appended)

ADPKD, autosomal dominant polycystic kidney disease; ARPKD, autosomal recessive polycystic kidney disease; JBS, Joubert syndrome; MKS, Meckel syndrome; SLS, Senior-Løken syndrome; BBS, Bardet-Biedl syndrome; ADTKD, autosomal dominant tubulointerstitial kidney disease; ADPLD, autosomal dominant polycystic liver disease

**Supplementary table S2. Target regions for adaptive sampling (T-LRS).**

| ***Gene*** | ***Target (hg38)*** | ***Target Size (bp)*** |
|---|---|---|
| *ARL13B* | chr3:93480139-94155678 | 675,539 |
| *CC2D2A* | chr4:14968660-15701971 | 733,311 |
| *IFT140* | chr16:1010427-1712072 | 701,645 |
| *NPHP1* | chr2:109622311-110305013 | 682,702 |
| *PKHD1* | chr6:51114685-52187625 | 1,072,940 |
| *TCTN2* | chr12:123171108-123808405 | 637,297 |

### Supplementary table S3. Clinical information on cases unresolved by panel screening

| No. | Pt. ID | Family ID | Category | UW-CRDR ID | Sex | Serum Cr (mg/dl) | eGFR (mL/min/1.73m2) | TKV (ml) | PLD | HT | Phenotype |
|---|---|---|---|---|---|---|---|---|---|---|---|
| 1 | 544 | K544 | 3 | USPKD1 | F | 0.28 | normal | N.A. | No | No | PKD |
| 2 | 545 | K545 | 3 | USPKD2 | M | 1.33 | 41.62 | 289 | Yes | Yes | PKD |
| 3 | 550 | K550 | 1 | USPKD3 | F | 0.82 | 60.52 | 334 | No | No | PKD |
| 4 | 563 | K563 | 3 | USPKD4 | M | ESKD | ESKD | 5681 | Yes | Yes | PKD |
| 5 | 564 | K564 | 3 | USPKD5 | F | 0.58 | 85.65 | 616 | No | Yes | PKD |
| 6 | 570 | K570 | 3 | USPKD6 | M | 1.05 | 53.48 | 587 | No | Yes | PKD |
| 7 | 590 | K590 | 3 | USPKD7 | F | ESKD | ESKD | N/A | N/A | N/A | MCKD |
| 8 | 602 | K602 | 2 | USPKD8 | M | 1.05 | 52.67 | 5182 | No | Yes | PKD |
| 9 | 619 | K619 | 3 | USPKD9 | M | ESKD | ESKD | 1415 | No | Yes | PKD |
| 10 | 629 | K629 | 3 | USPKD10 | F | 0.86 | 51.25 | 421 | No | No | PKD |
| 11 | 691 | K691 | 3 | USPKD11 | M | 0.75 | 89.13 | 1476 | No | Yes | PKD |
| 12 | 696 | K696 | 3 | USPKD12 | M | 0.91 | 71.68 | 328 | Yes | Yes | PKD |
| 13 | 697 | K697 | 2 | USPKD13 | F | 6.42 | 5.82 | 3628 | Yes | Yes | PKD |
| 14 | 698 | K698 | 3 | USPKD14 | M | 2.34 | 21.76 | 698 | No | Yes | PKD |
| 15 | 699 | K699 | 3 | USPKD15 | F | 1.13 | 38.92 | 1833 | No | Yes | PKD |
| 16 | 702 | K702 | 3 | USPKD16 | M | 2.14 | 23.83 | 238 | Yes | Yes | PKD |
| 17 | 730 | K730 | 2 | USPKD17 | M | 0.8 | 84.71 | N/A | No | Yes | PKD |
| 18 | 742 | K742 | 3 | USPKD18 | F | 2.12 | 23.10 | N/A | N/A | N/A | MCKD |
| 19 | 754 | K754 | 3 | USPKD19 | F | 0.71 | 67.09 | 1150 | No | Yes | PKD |
| 20 | 761 | K761 | 3 | USPKD20 | F | 1.11 | 40.49 | 1919 | No | Yes | PKD |
| 21 | 762 | K762 | 3 | USPKD21 | F | 0.65 | 76.07 | 2518 | Yes | No | PKD |
| 22 | 789 | K789 | 3 | USPKD22 | M | 1.34 | 43.49 | N/A | No | Yes | PKD |
| 23 | 835 | K835 | 3 | USPKD23 | M | 1.78 | 32.52 | 389 | No | Yes | PKD |
| 24 | 843 | K843 | 3 | USPKD24 | M | 1.02 | 57.55 | 4188 | No | Yes | PKD |
| 25 | 858 | K858 | 3 | USPKD25 | F | 0.51 | 94.82 | 225 | No | No | PKD |
| 26 | 868 | K868 | 1 | USPKD26 | M | ESKD | ESKD | 703 | No | Yes | PKD |
| 27 | 872 | K872 | 3 | USPKD27 | M | 1.32 | 44.21 | 304 | No | Yes | PKD |
| 28 | 875 | K875 | 3 | USPKD28 | F | 2.18 | 19.35 | N/A | N/A | N/A | MCKD |
| 29 | 891 | K891 | 3 | USPKD29 | F | 0.7 | 62.07 | N/A | No | Yes | PKD |
| 30 | 911 | K911 | 3 | USPKD30 | F | 0.94 | 48.32 | 231 | No | Yes | PKD |
| 31 | 913 | K913 | 3 | USPKD31 | F | 3.6 | 10.27 | N/A | N/A | N/A | MCKD |
| 32 | 914 | K914 | 3 | USPKD32 | F | 3.49 | 10.39 | N/A | N/A | N/A | MCKD |
| 33 | 942 | K942 | 3 | USPKD33 | M | 0.91 | 68.82 | 4359 | No | N/A | PKD |
| 34 | 955 | K955 | 3 | USPKD34 | M | ESKD | ESKD | 225 | No | Yes | PKD |
| 35 | 958 | K958 | 3 | USPKD35 | M | 1.2 | 51.42 | 568 | No | yes | PKD |
| 36 | 962 | K962 | 2 | USPKD36 | M | ESKD | ESKD | 3777 | No | Yes | PKD |
| 37 | 976 | K976 | 3 | USPKD37 | M | 8.66 | 5.82 | 1848 | No | Yes | PKD |
| 38 | 977 | K977 | 3 | USPKD38 | M | 11.28 | 4.59 | 11338 | Yes | Yes | PKD |
| 39 | 978 | K978 | 3 | USPKD39 | M | 3.39 | 15.91 | 207 | No | Yes | PKD |
| 40 | 993 | K993 | 2 | USPKD40 | M | 4.6 | 11.34 | 1193 | No | Yes | PKD |
| 41 | 1011 | K1011 | 3 | USPKD41 | M | ESKD | ESKD | 3495 | Yes | Yes | PKD |
| 42 | 1044 | K1044 | 3 | USPKD42 | F | 2.74 | 20.14 | N/A | N/A | N/A | MCKD |
| 43 | 1049 | K1049 | 2 | USPKD43 | F | 6.19 | 8.26 | N/A | N/A | N/A | MCKD |
| 44 | 1146 | K1146 | 3 | USPKD44 | M | 2.8 | 18.90 | 860 | No | Yes | PKD |
| 45 | 1158 | K1158 | 3 | USPKD45 | M | 1.05 | 56.52 | 1893 | No | Yes | PKD |
| 46 | 1171 | K1171 | 3 | USPKD46 | F | 6.62 | 5.3 | N/A | N/A | N/A | MCKD |
| 47 | 1192 | K1192 | 2 | USPKD47 | F | 2.36 | 19.17 | N/A | N/A | N/A | MCKD |
| 48 | 1206 | K1206 | 3 | USPKD48 | M | 0.91 | 68.82 | 1277 | No | Yes | PKD |
| 49 | 1216 | K1216 | 2 | USPKD49 | M | 2.72 | 20.24 | 720 | No | Yes | PKD |

TKV, Total Kidney Volume; PLD, Polycystic Liver Disease; HT, Hypertension; N/A, Not Available

**Supplementary table S4.**

**List of cases and variants profile in which the responsible mutation was identified by WGS or T-LRS in known genes**

| No. | Category | Pt._ID | UW-CRDR ID | Sex | Mutation in known genes | Impact | Zygosity | gnomAD (total) | 54KJPN | CADD | Splice AI | ACMG classification | Reports |
|---|---|---|---|---|---|---|---|---|---|---|---|---|---|
| 8 | 2 | 602 | USPKD8 | M | *IFT140* (NM_014714) c.1795dupA:p.Ile599AsnfsTer7 | frameshift | het. | N/A | N/A | N/A | − | Pathogenic [PVS1,PM2,PP5] | Schmidts M (2014) Hum Mutat. 34, 714 |
| 17 | 2 | 730 | USPKD17 | M | *IFT140* (NM_014714) c.2500C>T: p.Arg834* | nonsense | het. | 0.000007796 | N/A | 53 | − | Pathogenic [PVS1,PM2,PP5] | Schmidts M (2014) Hum Mutat. 34, 714 |
| 21 | 3 | 762 | USPKD21 | F | *PKD2* (NM_000297.3) c.1550_1553dup:p.Val520SerfsTer7 | frameshift | het. | N/A | N/A | N/A | − | Likely-pathogenic [PVS,PM2] | novel |
| 37 | 3 | 976 | USPKD37 | M | *PKD1* (NM_001009944) c.8017-2_8017-1del | splicing | het. | 0.000001324 | N/A | N/A | Splice-Altering / strong (0.96) | Pathogenic [PVS1,PM2,PS4, PS3,PP5] | Rossetti S (2000) Am J Hum Genet 68: 46 |
| 38 | 3 | 977 | USPKD38 | M | *PKD1* (NM_001009944) c.9564_9566del:p.Asn3188del | in-frame deletion | het. | N/A | N/A | N/A | − | Pathogenic [PS4,PM2,PM4, PM5,PM1,PP1,PP5] | novel |
| 43 | 2 | 1049 | USPKD43 | F | *NPHP1* c.1639C>T:p.Gln547* (P) | nonsense | het. | N/A | 0.0001 | 40 | − | Likely-Pathogenic [PVS1,PM2] | Fujimaru (2021) Kidney Int Rep 6, 1346 |
| | | | | | 7,401 bp deletion in NPHP1 | deletion | het. | N/A | N/A | N/A | − | N/A | none |
| 45 | 3 | 1158 | USPKD45 | M | *IFT140* (NM_014714) c.2068-2A>G | splicing | het. | 0.000004541 | N/A | N/A | Splice-Altering / strong (0.99) | Pathogenic [PVS1,PM3,PM2,PP5] | novel |
| 47 | 2 | 1192 | USPKD47 | F | *NPHP1* 80 kb deletion | deletion | het. | N/A | N/A | N/A | − | N/A | novel |
| | | | | | *NPHP1* 207 bp deletioin | deletion | het. | N/A | N/A | N/A | − | N/A | novel |

**Supplementary Information References**


1. Mori T, Hosomichi K, Chiga M*, et al.* Comprehensive genetic testing approach for major inherited kidney diseases, using next-generation sequencing with a custom panel. *Clin Exp Nephrol* 2017; **21:** 63-75.

2. Fujimaru T, Mori T, Sekine A*, et al.* Kidney enlargement and multiple liver cyst formation implicate mutations in PKD1/2 in adult sporadic polycystic kidney disease. *Clin Genet* 2018; **94:** 125-131.

3. Fujimaru T, Kawanishi K, Mori T*, et al.* Genetic Background and Clinicopathologic Features of Adult-onset Nephronophthisis. *Kidney Int Rep* 2021; **6:** 1346-1354.

4. Li H, Durbin R. Fast and accurate short read alignment with Burrows-Wheeler transform. *Bioinformatics* 2009; **25:** 1754-1760.

5. Li H, Handsaker B, Wysoker A*, et al.* The Sequence Alignment/Map format and SAMtools. *Bioinformatics* 2009; **25:** 2078-2079.

6. Van der Auwera GA, Carneiro MO, Hartl C*, et al.* From FastQ data to high confidence variant calls: the Genome Analysis Toolkit best practices pipeline. *Curr Protoc Bioinformatics* 2013; **43:** 11.10.11-11.10.33.

7. Chong JX, Buckingham KJ, Jhangiani SN*, et al.* The Genetic Basis of Mendelian Phenotypes: Discoveries, Challenges, and Opportunities. *Am J Hum Genet* 2015; **97:** 199-215.

8. Chen S, Francioli LC, Goodrich JK*, et al.* A genomic mutational constraint map using variation in 76,156 human genomes. *Nature* 2024; **625:** 92-100.

9. Paila U, Chapman BA, Kirchner R*, et al.* GEMINI: integrative exploration of genetic variation and genome annotations. *PLoS Comput Biol* 2013; **9:** e1003153.

10. Kircher M, Witten DM, Jain P*, et al.* A general framework for estimating the relative pathogenicity of human genetic variants. *Nat Genet* 2014; **46:** 310-315.

11. Kumar P, Henikoff S, Ng PC. Predicting the effects of coding non-synonymous variants on protein function using the SIFT algorithm. *Nat Protoc* 2009; **4:** 1073-1081.

12. Adzhubei I, Jordan DM, Sunyaev SR. Predicting functional effect of human missense

mutations using PolyPhen-2. *Curr Protoc Hum Genet* 2013; **Chapter 7:** Unit7.20.

13. Jagadeesh KA, Wenger AM, Berger MJ*, et al.* M-CAP eliminates a majority of variants of uncertain significance in clinical exomes at high sensitivity. *Nat Genet* 2016; **48:** 1581-1586.

14. Davydov EV, Goode DL, Sirota M*, et al.* Identifying a high fraction of the human genome to be under selective constraint using GERP++. *PLoS Comput Biol* 2010; **6:** e1001025.

15. Pais LS, Snow H, Weisburd B*, et al.* seqr: A web-based analysis and collaboration tool for rare disease genomics. *Hum Mutat* 2022; **43:** 698-707.
